# Divergent trends in the incidence and mortality of coronary events, especially in women. Evidence from Finland in 1996-2021

**DOI:** 10.1101/2023.12.01.23299126

**Authors:** Atte Kallström, Ida Holopainen, Oleg Kambur, Markus Perola, Veikko Salomaa, Aki S. Havulinna, Markus Ramste, Juha Sinisalo

## Abstract

**Objective:** Acute coronary syndrome (ACS) incidence and case fatality (CF) have declined in the past decades, but some studies have suggested a potential stagnation in this decline. We examined how the ACS burden has evolved among persons aged 35-74 in Finland from 1996 to 2021.

**Methods:** We used Finnish country-wide Hospital Discharge- and Causes of Death-Registers covering first non-fatal and fatal ACS events, totaling 69 906 442 person-years at risk. We analyzed incidence, mortality, and 28-day CF, and their trends using negative binomial, Poisson, segmented, and logistic regression adjusting for age and sex.

**Results:** Altogether, the analysis consisted of 186 489 non-fatal and 72 907 fatal ACS events. ACS incidence declined in men (annual percentage change (APC) −2.0% [95% CI −2.2 to −1.8]) and in older women (APC of 55-64 year old −1.5% [−1.7 to −1.2] and 65-74 year old −3.3% [−3.4 to −3.2]), but the incidence decline slowed down over the last decade. In younger women aged 35-54, incidence was unchanged during the study period. ACS mortality and CF declined (APC of the mortality in men - 4.4% [−4.6 to −4.2] and in women −5.0% [−5.2 to −4.7]. APC of CF in men −2.7% [−2.8 to −2.6] and in women −3.3% [−3.6 to −3.1]).

**Conclusions:** ACS mortality declined in all groups, but the decline in ACS incidence slowed down and even halted in women. In women aged 35-54, the incidence was unchanged during the study period. These results emphasize the need of intensified cardiovascular prevention, particularly in women.

**Key messages:** *What is already known on this topic:* *During the last decades coronary artery disease treatment and prevention have improved worldwide which has led to a decline in ACS mortality and case fatality. However, recent studies from several countries suggest, that incidence decline has stagnated or even turned to increase, especially in younger age cohorts.*

*What this study adds:* *This study showed that incidence has ceased its decline in women aged 35-54 and has slowed down in older age groups. Together with declining mortality this results in a growing number of patients living with cardiovascular disease leading to increased healthcare costs*.

*How this study might affect research, practice, or policy:* *The alarming results of this study underline the importance of intensifying prevention, focusing especially on young and middle-aged women*.

## Introduction

Cardiovascular disease is one of the world’s leading causes of death and has been extensively researched ^1^ ^2^. Established modifiable risk factors are hypertension, dyslipidemia, diabetes mellitus, and smoking. Preventive measures that target these risk factors have been shown to be effective and have a good cost-benefit ratio ^3^. With these measures, acute coronary syndrome (ACS) mortality rates have declined during the past decades in Western countries. Approximately one-third of total ACS mortality decline is due to an increase in survival after myocardial infarction ^4^. Compared to earlier decades, patients now have fewer and less severe symptoms, and survival rates are higher. However, there are worrying signs that these positive trends may not be sustainable, as shown by data from the US and Australia. ^5^ ^6^. It has been proposed that the alarming increases in obesity and diabetes mellitus worldwide are reflected in ACS incidence ^5^ ^7^.

In Finland, the coronary heart disease mortality rate has drastically declined during the preceding decades. In the last century from 1960 to 1970 Finland’s mortality rate from ACS was among the highest in the world. Most of the reduction is due to improving primary and secondary prevention, especially concentrating on the classical cardiovascular risk factors. ^8^ Also, a substantial part of the reduction has been gained by advances in the treatment of ACS, including percutaneous coronary intervention and new preventive medications ^9^. Still, the northeastern part of Finland has a substantially higher burden of CHD incidence and mortality, partly due to genetic, environmental, and socioeconomic aspects ^8^ ^10^. Previous studies have demonstrated that the short-term case fatality and 1-year prognosis of incident MI have improved in both sexes from the mid-1990s to 2002 in Finland ^11^.

Data on trends in ACS incidence and mortality rates, and case fatality during the last years is very sparse all over the world as well as in Finland ^8^ ^12^. As the Finnish electronic health care registries offer reliable and comprehensive nationwide data and a long follow-up period ^13^, we chose to analyze the trends using these registries and compare our findings with available corresponding data from other Western countries. We focused on ACS trends in the 35-74 year old population of Finland from 1996 to 2021. We hypothesized that these trends would be declining since ACS mortality in the Western world has declined in the past and it was previously projected that age-adjusted ACS event rates would decrease in Finland until 2050 ^14^.

## Methods

### Study population

We collected data from Finnish country-wide registers, i.e., hospital discharges, from the Care Register for Health Care and the National Causes of Death Register with complete coverage of non-fatal and fatal ACS events, including sudden, out-of-hospital cardiac deaths. A unique personal identification number that every permanent resident in Finland receives at birth or upon immigration links the data between the registries. Altogether, the analysis consists of 186 489 non-fatal and 72 907 fatal incident ACS events.

We defined annual population as the population on the last day of each year, available from the Finnish Population Information System. At the end of 2021, the population of Finland aged 35-74 was 2.8 million. The study population contributed 69 906 442 person-years at risk.

The information permit number for the use of the Finnish Cardiovascular Disease Register (CVDR) is THL/3624/6.02.00/2023. Permission to access the original data can be applied for through FINDATA.

### Definition of ACS events

The data contains only the first non-fatal and fatal ACS events, as previously classified in the WHO-MONICA project ^15^. We defined the event as first (i.e., incident) for an individual if he/she had no previous ACS events recorded in the HDR during the preceding 10 years. If a patient had a second ACS event less than 28 days from the onset of the previous event, we considered two events as one and considered only the more severe diagnosis code. We defined an ACS event fatal if the patients died within 28 days from the onset of the event and CF as the proportion of fatal cases of all incident cases.

Total ACS events include non-fatal hospitalizations for MI or unstable angina (ICD-9: 410; ICD-10: I20.0, I21-I22), and fatal events, which include deaths with CHD (ICD-9: 410-414; ICD-10: I20-I25), cardiac arrest (ICD-10: I46), sudden death for unknown reason (Finnish ICD-9: 798, not 7980A; ICD-10: R96), or unwitnessed death (ICD-10: R98) as the underlying or direct cause of death, or deaths with MI (ICD-9: 410; ICD-10: I21-I22) as the contributing cause of death.

We assessed ACS incidence, mortality, and case fatality for the entire population of Finland by event age, and by event year and sex. Experimental setup is shown as flowchart in Supplemental Figure 2.

### Age groups

We calculated ACS incidence, mortality, and case fatality for the age group 35-74 and further subdivided into smaller age groups as follows 35-44, 45-54, 55-64, and 65-74. CF was an exception since the two youngest age groups were combined into one 35-54 age group to lower the random variation due to low case counts.

### Statistics

We analyzed the incidence and mortality rates, as well as the significance of their trends using Poisson or negative binomial (NB) regression models adjusting for age and sex. We used NB models instead of the Poisson model if there was evidence of overdispersion, and the NB model had a better fit for the given data. We analyzed case fatality trends using the logistic regression model.

We used the segmented regression model to analyze the ACS incidence, mortality, and CF trend changes during the study period.

We performed age-standardization of the incidence and mortality rates using the direct method with weights from the 2011-2030 European standard population^16^. This standard population allows further comparison with other publications. The weights used for age-standardizing case fatality are based on the age distribution of observed coronary events in MONICA populations in the WHO-MONICA project ^15^. However, these weights were not defined for older age groups, thus we used weights aimed at a larger age distribution and smaller age groups (Supplemental Table 2).

The ACS incidence and mortality rates as well as case-fatality trends are presented in the figures as three-year moving averages, the first and the last years are counted as two-year averages.

We used R version 4.2.2 for all statistical analyses and considered p<0.05 as statistically significant. All tests were two-tailed. We used package “MASS” (version 7.3-58.1) for negative binomial regression, “segmented” (version 1.6-2) for segmented regression, and “AER” (version 1.2-10) for testing whether there was overdispersion in Poisson models. ^17^ The code used for the data analysis is available upon reasonable request.

## Results

### Overall incidence and mortality

In men and women, ACS incidence and mortality declined overall during the study period. (Figure 1). The decline in mortality was markedly steeper than the decline in incidence. The age-standardized ACS mortality rate declined by more than two thirds (in men 65.3% and in women 71.8%) and the incidence rate declined approximately by one third in men (35.1%) and by one quarter in women (24.6%).

**Figure 1:**
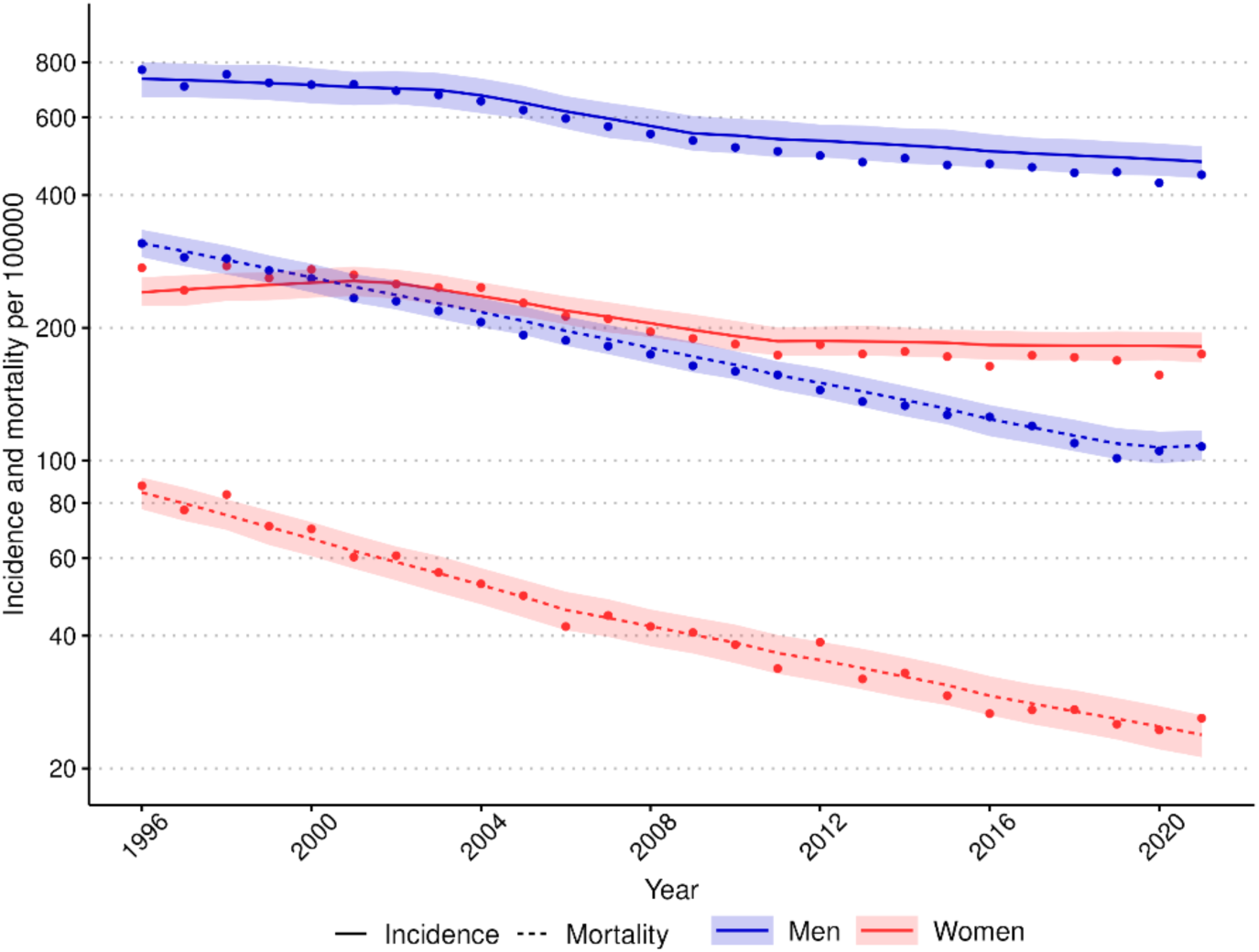
Trends in incidence and mortality rates of ACS in men and women aged 35-75 years, 1996-2021. Rates include first non-fatal I20.0, I21, and I22 cases and fatal I20-25, I46, R96 and, R98 cases. Age-standardized rates per 100 000 inhabitants were calculated with the 2013 European standard population as the reference. Observed incidence and mortality rates are presented as dots, the segmented (negative binomial) regression model’s predicted values as a line and the segmented regression model’s 95%-confidence intervals for predictions as a ribbon. The rates are presented on a logarithmic scale.

Men have approximately two times higher incidence than women in the oldest age group and four times higher incidence in younger age groups (35-54 year old) (Figure 2A&B). Men have also approximately three times higher mortality in the oldest age group (65-74 year old) and five times higher mortality rate in the other age groups (Figure 2C&D). Annual incidence and mortality changes per age group, number of cases in the age groups, mean age of each group, and p-values of trends can be seen in Table 1.

**Figure 2:**
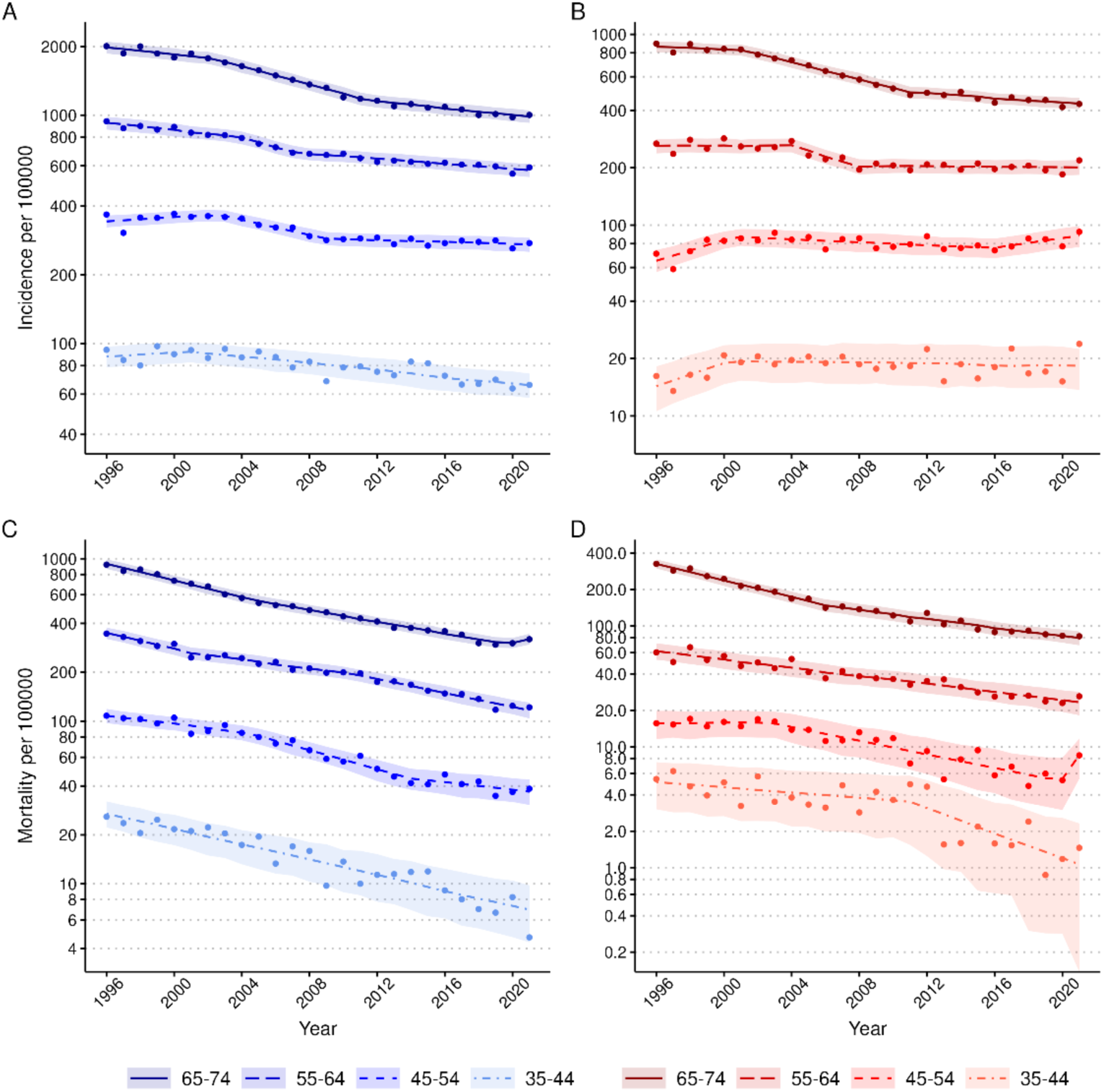
Trends in incidence and mortality rates of ACS in men and women by age group, 1996-2021. A) Incidence rates of men. B) Incidence rates of women. C) Mortality rates of men. D) Mortality rates of women. Incidence rates include first non-fatal I20.0, I21, and I22 and fatal I20-25, I46, R96, and R98 cases. Mortality rates include the fatal cases as mentioned above. Age-standardized rates per 100 000 inhabitants were calculated with the 2011-2030 European standard population as the reference. Observed incidence and mortality rates are presented as dots, the segmented, Poisson or negative binomial regression model’s predicted values as a line, and the regression model’s 95%-confidence intervals for predictions as a ribbon. The rates are presented on a logarithmic scale.

**Table 1:** Average annual changes in acute coronary syndrome incidence, mortality, and case fatality (CF) from 1996 to 2021. Incidence and mortality rate trends were analyzed using a Poisson or negative binomial regression model. CF trends were analyzed with a logistic regression model. The rate changes are presented as average annual change percentages during the study period. P-values represent the significance of the trend by study year. *Negative binomial model was used instead of Poisson model.

| Incidence |  |  |  |  |  |
| --- | --- | --- | --- | --- | --- |
| Age group | Mean age | Cases (N) | Average annual change (%) | 95% CI | P-value |
| **Men** |  |  |  |  |  |
| All (35-74)* | 62.0 | 185 309 | -2.01 | -2.20 to -1.82 | <1e-16 |
| 65-74* | 69.6 | 84 763 | -3.12 | -3.29 to -2.96 | <1e-16 |
| 55-64* | 59.9 | 61 912 | -2.06 | -2.23 to -1.89 | <1e-16 |
| 45-54* | 50.4 | 30 985 | -1.40 | -1.57 to -1.22 | <1e-16 |
| 35-44 | 40.9 | 7 649 | -1.33 | -1.63 to -1.04 | <1e-16 |
| **Women** |  |  |  |  |  |
| All (35-74)* | 64.7 | 74 087 | -1.66 | -1.83 to -1.48 | <1e-16 |
| 65-74* | 70.1 | 44 294 | -3.28 | -3.40 to -3.16 | <1e-16 |
| 55-64* | 60.2 | 20 304 | -1.46 | -1.70 to -1.21 | <1e-16 |
| 45-54 | 50.5 | 7 818 | 0.20 | -0.10 to 0.50 | 0.19 |
| 35-44 | 41.0 | 1 671 | 0.38 | -0.25 to 1.02 | 0.24 |
| Mortality |  |  |  |  |  |
| **Men** |  |  |  |  |  |
| All (35-74)* | 64.5 | 56 900 | -4.42 | -4.59 to -4.24 | <1e-16 |
| 65-74* | 69.6 | 30 602 | -4.49 | -4.68 to -4.30 | <1e-16 |
| 55-64* | 60.4 | 18 028 | -4.05 | -4.29 to -3.82 | <1e-16 |
| 45-54 | 50.8 | 6 835 | -4.77 | -5.09 to -4.45 | <1e-16 |
| 35-44 | 40.7 | 1 435 | -5.29 | -5.98 to -4.60 | <1e-16 |
| **Women** |  |  |  |  |  |
| All (35-74)* | 66.2 | 16 007 | -5.00 | -5.21 to -4.72 | <1e-16 |
| 65-74* | 70.1 | 11 104 | -5.50 | -5.78 to -5.21 | <1e-16 |
| 55-64 | 60.6 | 3 488 | -3.84 | -4.28 to -3.39 | <1e-16 |
| 45-54 | 50.7 | 1 104 | -4.47 | -5.27 to -3.67 | <1e-16 |
| 35-44 | 40.9 | 311 | -4.87 | -6.36 to -3.40 | 3.06e-10 |
| Case fatality |  |  |  |  |  |
| **Men** |  |  |  |  |  |
| All (35-74) | 62.0 | 185 309 | -2.71 | -2.84 to -2.57 | <1e-16 |
| 65-74 | 69.6 | 84 763 | -2.19 | -2.37 to -2.01 | <1e-16 |
| 55-64 | 59.9 | 61 912 | -2.75 | -3.00 to -2.52 | <1e-16 |
| 35-54 | 48.5 | 38 634 | -4.45 | -4.78 to -4.12 | <1e-16 |
| **Women** |  |  |  |  |  |
| All (35-74) | 64.7 | 74 087 | -3.35 | -3.58 to -3.12 | <1e-16 |
| 65-74 | 70.1 | 44 294 | -3.15 | -3.43 to -2.88 | <1e-16 |
| 55-64 | 60.2 | 20 304 | -2.79 | -3.28 to -2.30 | <1e-16 |
| 35-54 | 48.9 | 9 489 | -5.74 | -6.51 to -5.00 | <1e-16 |

### Incidence trends

The trends in ACS incidence are presented in Figure 2 A) men and B) women. In men, the incidence of ACS declined significantly in all age groups during the study period (annual percentage change (APC) in men −2.0% [95% CI −2.2 to −1.8]) (Table 1). Segmented regression however revealed that the incidence decline has slowed down significantly overall in men and men’s age groups 65-74, and 55-64 and halted in age group 45-54 from around 2007-2012 onwards (Supplemental Table 1, Supplemental Figures 3&4A).

In women, the incidence rate declined significantly overall and in the older age groups (overall APC −1.7 [−1.8; −1.5], among 65-74 year old −3.3% [−3.4 to −3.2]) and among 55-64 year old −1.5% [−1.7 to −1.2]). The overall incidence in women did not decline after 2010 and age-specific incidence rates slowed down in the older age groups from 2008-2011 (Supplemental Table 1, Supplemental Figures 3&4B). Importantly, in women aged 35-54, ACS incidence stayed the same for over two decades (APC of 35-44 year old 0.4% [−0.3 to 1.0] and 45-54 year old 0.2% [−0.1 to 0.5]). The stagnated or slowed incidence decline is mostly associated with inferior development of non-fatal incident ACS events compared to the incidence of all ACS events (Supplemental Figure 1A-C and Supplemental Table 3). The incidence of non-fatal ACS events among young women aged 35-54 has increased during the study period.

### Mortality trends

The trends in ACS mortality are presented in Figure 2 C) men and D) women and the breakpoints in Supplemental Figure 4C-D. The ACS mortality declined in both sexes and all age groups during the study period (APC of the mortality in men - 4.4% [−4.6 to −4.2] and in women −5.0% [−5.2 to −4.7]). Overall, the annual mortality change was very similar in all age groups. In women, mortality declined most in the oldest age group (APC −5.5% [−5.8 to −5.2]) while in men, mortality declined most in the youngest age group (APC −5.3% [−6.0 to −4.6]). However, in younger age groups, especially in women, case numbers are low (Table 1).

### 28-day case-fatality trends

ACS case-fatality declined over the study period in men and women (APC of the CF in men −2.7% [−2.8 to −2.6] and in women −3.3% [−3.6 to −3.1]) (Figure 3A). Women exhibited lower ACS case-fatality rates across equivalent age groups compared to men (Figure 3B). The decline was not linear, particularly in the older age groups. The case fatality stayed the same or even increased a little from around 2001 to 2010, depending on age group and sex (Supplemental Table 1 and Supplemental Figure 5A&B). The annual decline in case fatality was most significant in the youngest age group, almost two times faster in women and almost as fast in men compared to the older age groups.

**Figure 3:**
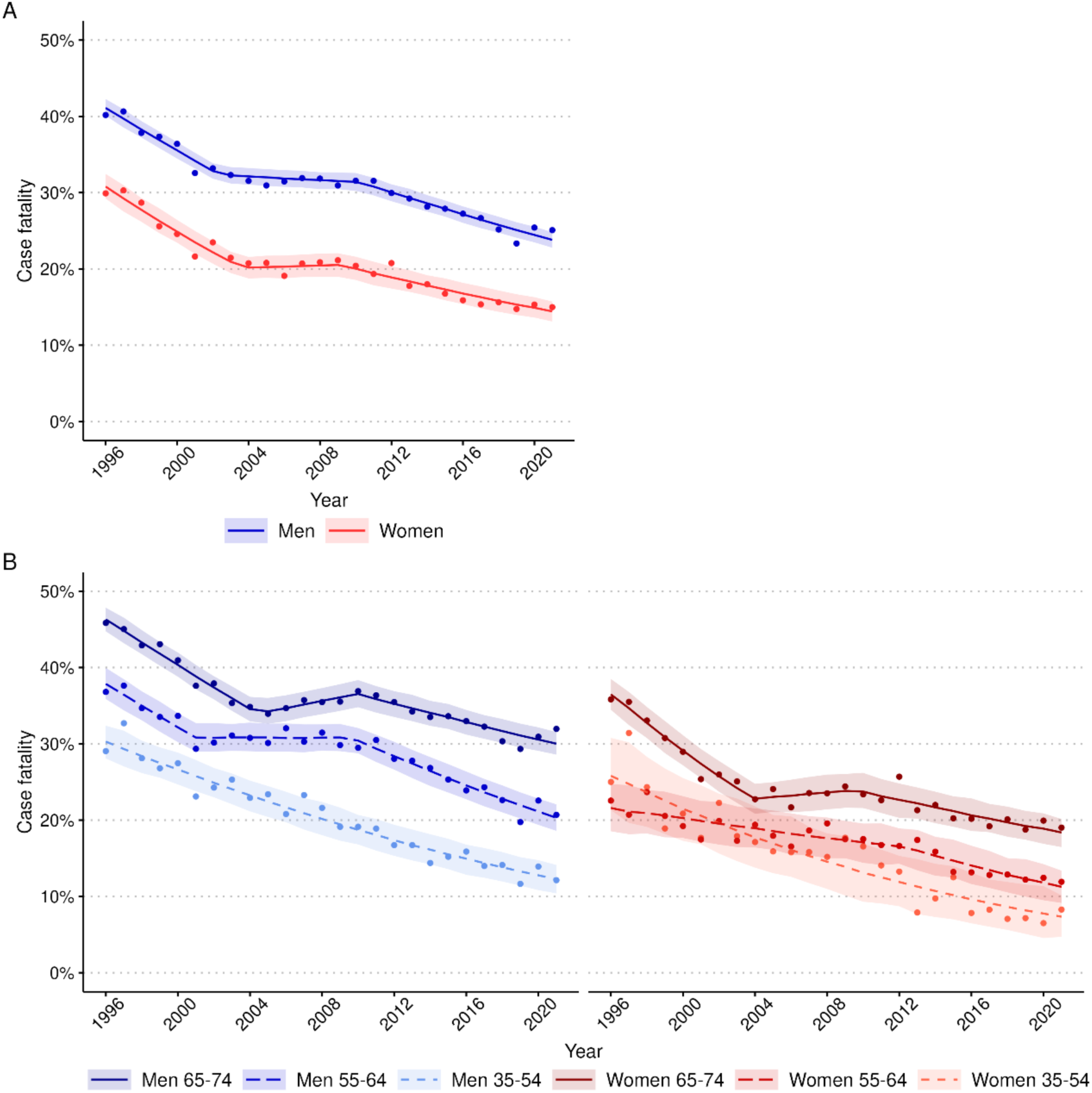
Trends in case fatality (CF) of acute coronary syndrome (A) among men and women aged 35-74, and (B) by 10-year age group (B), 1996-2021. CF includes the first non-fatal I20.0, I21, and I22 and fatal I20-25, I46, R96, and R98 cases. CF was age-standardized using weights based on the age distribution of observed coronary events in populations participating in the WHO-MONICA project. Observed case-fatality is presented as dots, the segmented or logistic regression model’s predicted values as a line and the regression model’s 95%-confidence intervals for predictions as a ribbon

## Discussion

In this nationwide study of almost 260 000 cases, we found a declining incidence of acute coronary syndrome (ACS) in the population aged 35-74 during the study period of 25 years. In young women aged 35-54, however, the incidence did not decline. Also, in the oldest age groups, the declining incidence trends were slowed down during the study period. Furthermore, the overall incidence decline in women halted in the last decade. Other reports have shown that the ACS incidence trend has turned into increase, especially among younger age groups ^5^ ^18^. Our study also showed that the mortality rate declined in men and women and, unlike incidence, in all age groups throughout the study period. The rate of decline was approximately similar in all age groups. The decline in mortality is in line with the earlier prediction ^14^. It is also worth noting that the decline in mortality was much faster than in incidence. Case fatality declined similarly to mortality in both men and women and in all age groups during the study period. The decline in case fatality was most significant in the youngest age group which consisted of people aged 35-54 years. The case fatality was overall much lower in women than in men.

To understand why the incidence of ACS is no longer steadily declining, we examined the results of the National FinHealth 2017 survey that has gathered information on the health of the Finnish adult population and on the risk factors influencing their health ^19^. The FinHealth study showed that the proportion of smokers has declined. The study also showed that the prevalence of elevated LDL has declined, but no changes in low HDL levels were seen. Alcohol abstinence has also increased. However, dietary habits have not improved, obesity has increased, as well as the waist-to-hip ratio, and hypertension is still very common in Finland. In summary, except for obesity and diabetes, other ACS-related risk factors have improved.

One explanation for the halted incidence decline in women aged 35-54 could be that diabetes is a stronger risk factor in women than in men ^20^, considering that diabetes prevalence has also increased ^19 21^. Also, even as smoking has lost popularity, tobacco exposure, both current and accumulated, predisposes female smokers to more premature myocardial infarction than men ^22^. Additionally, the use of oral contraceptives, which has increased in popularity in the last decades, could have influenced this negative development. However, the literature regarding their effect on the risk of ischemic heart disease is unanimous ^23–25^. In addition to these somatic factors, psychosocial factors may play a role. It has been shown that women under 50 years of age who develop a myocardial infarction have more psychosocial risk factors than same-aged men and depressive symptoms have been shown to be associated with chronic heart disease events ^26^.

Regarding prevention, studies have shown that primary prevention is implemented even better in women than in men, but women receive less invasive measures and guideline suggested secondary preventive medication than men after myocardial infarction. Although women have a lower risk for recurrent myocardial infarction and lower numbers of 30-day cardiovascular deaths compared to men, women are reported to have a higher rate of long-term all-cause mortality.^27^

In the older age groups, the slowing down of the incidence decline may be due to the exhaustion in optimizing preventive treatment and therapeutic inertia, especially when it comes to incorporating new expensive treatment modalities (e.g., PCSK9, siRNA drugs) that will be utilized in younger age groups ^28^. In addition, for all age groups, improving ACS prevention and reducing mortality beyond optimal treatment of classical risk factors may require a novel avenue of targeting the residual risk that may not be related to classical risk factors but mediated through inflammation and other poorly understood pathomechanisms within the vascular wall ^30^. Recently new tools, such as polygenic risk scores, have been developed to predict the residual and genetic risk of ACS ^31^.

A factor that could have an overall influence on the development of incidence and case fatality in the 21st century is the introduction of sensitive troponins to clinical practice, which have the power to detect smaller myocardial infarctions.^32^. Additionally, the COVID-19 pandemic may have influenced the incidence and mortality figures of the last two years as COVID-19 is a risk factor for acute coronary syndrome ^33^.

In concordance with other recent reports ^34^ ^35^, we showed that ACS mortality declined steadily in women and men in all age groups. Awareness of coronary artery disease might influence earlier seeking of care, which results in lower mortality. Because of this favorable trend in mortality, Finland is included in the moderate-risk region in Europe based on the death risk due to cardiovascular disease, leaving still room for improvement as many countries in Western Europe belong to the low-risk region ^36^. Taking this into account, a decline in mortality and incidence could be continuously achieved and improved, especially if the treatment methods evolve and there will be more focus on primary and secondary prevention.

Our results showed that case fatality declined rapidly in men and women, especially in the youngest age group. Finland’s positive trend might be explained by the same factors as the decrease in mortality. This can also be influenced by the introduction of sensitive troponins ^32^. In addition, we identified a plateau phase in the overall declining case fatality trend from 2002 to 2010. An explanation for this plateau phase in CF is not known.

Our study is based on comprehensive nationwide Finnish Cardiovascular Disease Register data which covers all symptomatic acute coronary syndrome incidents in Finland with a long study period of twenty-five years. However, the study should be interpreted with certain limitations in mind: the underlying Finnish health care registers do not provide individual information for example ECGs or troponin levels, thus the diagnoses of the cases cannot be further verified. Additionally, silent myocardial infarctions are not found in the registers. However, cardiovascular diagnoses in Finnish healthcare registers have been validated and found reliable which increases the confidence in our results ^13^.

## Conclusions

Our study demonstrated diverging trends in different components of ACS event rates in the Finnish population aged 35-74 years. The ACS mortality and CF are steeply declining, whereas the incidence decline is leveling off. Notably, the decline in incidence in older age groups has slowed during the last decade. Also, the overall incidence in women ceased to decline. Indirectly, this may result in an increase in CAD patients in the Finnish population. Consequently, the need for treatment, secondary prevention, and rehabilitation resources is likely to increase as well. Particularly alarming is the halted incidence decline in women aged 35-54 years. Since the incidence of first events can be reduced by primary prevention only, this emphasizes the importance of improving the primary prevention measures, focusing especially on young and middle-aged women.

## Data Availability

All code produced in the present study are available upon reasonable request to the authors. Permission to access the original data can be applied for through FINDATA.

## Author contributions

A.K., M.R., V.S., A.H. and J.S. wrote the manuscript. M.P., A.H., M.R, V.S., and J.S. designed the research. A.K., M.R., V.S., A.H. and J.S. performed the research. A.K., I.H., O.K. and A.H. analyzed the data.

## Acknowledgement

The authors wish to acknowledge CSC – IT Center for Science, Finland, for computational resources.

## Supplemental Material

**Supplemental Table 1:** Segmented (Poisson or negative binomial) regression model’s breakpoints and intervals of incidence, mortality, and case fatality trends. Intervals where there was no trend change are shown on a yellow background and increasing trends are shown on a red background.

| Incidence |  |  |  |  |
| --- | --- | --- | --- | --- |
| Age group | Breakpoints | Interval 1 (estimate + slope [95% CI]) | Interval 2 (estimate + slope [95% CI]) | Interval 3 (estimate + slope [95% CI]) |
| **Men** |  |  |  |  |
| All (35-74) | 2003, 2009 | -10.398 - 0.009 [-0.020; 0.002] * year | -10.175 - 0.039 [-0.062; -0.017] * year | -10.521 - 0.013 [-0.018; -0.007] * year |
| 65-74 | 2002, 2012 | -8.383 - 0.018 [-0.029; -0.007] * year | -8.215 - 0.045 [-0.053; -0.038] * year | -8.636 - 0.018 [-0.025; -0.012] * year |
| 55-64 | 2004, 2007 | -8.880 - 0.018 [-0.027; -0.009] * year | -8.662 - 0.047 [-0.073; -0.020] * year | -9.045 - 0.013 [-0.017; -0.009] * year |
| 45-54 | 2003, 2009 | -10.924 - 0.006 [-0.004; 0.017] * year | -10.626 - 0.037 [-0.049; -0.027] * year | -11.052 - 0.005 [-0.010; 0.000] * year |
| 35-44 | 2001 | -13.973 + 0.010 [-0.015; 0.034] * year | -13.840 - 0.017 [-0.021; -0.012] * year | - |
| **Women** |  |  |  |  |
| All (35-74) | 2001, 2011 | -12.619 + 0.012 [-0.004; 0.028] * year | -12.367 - 0.035 [-0.043; -0.026] * year | -12.810 - 0.004 [-0.011; 0.003] * year |
| 65-74 | 2001, 2011 | -11.582 - 0.008 [-0.022; 0.006] * year | -11.360 - 0.051 [-0.059; -0.043] * year | -11.888 - 0.015 [-0.021; -0.011] * year |
| 55-64 | 2004, 2008 | -12.083 + 0.001 [-0.010; 0.011] * year | -11.567 - 0.064 [-0.120; -0.008] * year | -12.315 - 0.001 [-0.007; 0.004] * year |
| 45-54 | 2001, 2016 | -13.431 + 0.060 [0.024; 0.097] * year | -13.121 - 0.009 [-0.015; -0.002] * year | -13.881 + 0.029 [-0.007; 0.066] * year |
| 35-44 | 2000 | -16.447 + 0.069 [-0.009; 0.147] * year | -16.132 - 0.003 [-0.011; 0.006] * year | - |
| Mortality |  |  |  |  |
| **Men** |  |  |  |  |
| All (35-74) | 2020 | -12.377 - 0.046 [-0.048; -0.044] * year | -13.618 + 0.007 [-0.101; 0.114] * year | - |
| 65-74 | 2005, 2020 | -10.349 - 0.060 [-0.068; -0.051] * year | -10.504 - 0.042 [-0.046; -0.037] * year | -12.784 + 0.054 [-0.050; 0.016] * year |
| 55-64 | 2001, 2010 | -11.605 - 0.055 [-0.075; -0.036] * year | -11.720 - 0.032 [-0.043; -0.021] * year | -11.477 - 0.049 [-0.058; -0.040] * year |
| 45-54 | 2005, 2014 | -13.462 - 0.033 [-0.046; -0.020] * year | -13.170 - 0.067 [-0.083; -0.050] * year | -13.895 - 0.027 [-0.052; -0.001] * year |
| 35-44 | - | - | - | - |
| **Women** |  |  |  |  |
| All (35-74) | 2006 | -15.091 - 0.061 [-0.069; -0.053] * year | -15.258 - 0.044 [-0.050; -0.038] * year | - |
| 65-74 | 2006 | -14.738 - 0.079 [-0.088; -0.069] * year | -15.106 - 0.042 [-0.049; -0.035] * year | - |
| 55-64 | - | - | - | - |
| 45-54 | 2003, 2020 | -14.752 - 0.001 [-0.049; 0.046] * year | -14.320 - 0.065 [-0.082; -0.048] * year | -26.779 + 0.456 [-0.136; 1.048] * year |
| 35-44 | 2011 | -15.841 - 0.025 [-0.055; 0.005] * year | -14.442 - 0.119 [-0.192; 0.045] * year | - |
| Case fatality |  |  |  |  |
| **Men** |  |  |  |  |
| All (35-74) | 2002, 2010 | -2.557 - 0.060 [-0.069; -0.052] * year | -2.908 - 0.006 [-0.013; 0.002] * year | -2.473 - 0.036 [-0.041; -0.031] * year |
| 65-74 | 2004, 2010 | -2.059 - 0.061 [-0.070; -0.053] * year | -2.746 + 0.020 [0.002; 0.038] * year | -2.088 - 0.027 [-0.034; -0.020] * year |
| 55-64 | 2001, 2010 | -2.917 - 0.062 [-0.083; -0.042] * year | -3.228 - 0.000 [-0.013; 0.013] * year | -2.561 - 0.049 [-0.057; -0.041] * year |
| 35-54 | - | - | - | - |
| **Women** |  |  |  |  |
| All (35-74) | 2004, 2009 | -2.881 - 0.075 [-0.088; -0.063] * year | -3.491 + 0.004 [-0.017; 0.026] * year | -2.959 - 0.036 [-0.045; -0.028] * year |
| 65-74 | 2004, 2010 | -3.380 - 0.085 [-0.099; -0.070] * year | -4.142 + 0.012 [-0.016; 0.04] * year | -3.582 - 0.029 [-0.040; -0.019] * year |
| 55-64 | 2012 | -3.724 - 0.021 [-0.030; -0.012] * year | -3.244 - 0.050 [-0.077; -0.023] * year | - |
| 35-54 | - | - | - | - |

**Supplemental Table 2:** The weights for age-standardizing case fatality. Based on the age distribution of observed coronary events in MNICA populations in the WHO-MONICA project.

| Age (years) | Weights |
| --- | --- |
| 0-4 | 1 |
| 5-9 | 1 |
| 10-14 | 1 |
| 15-19 | 1 |
| 20-24 | 1 |
| 25-29 | 1 |
| 30-34 | 2 |
| 35-39 | 5 |
| 40-44 | 9 |
| 45-49 | 16 |
| 50-54 | 26 |
| 55-59 | 42 |
| 60-64 | 56 |
| 65-69 | 75 |
| 70-74 | 93 |
| 75-79 | 100 |
| 80-84 | 100 |
| 85- | 100 |

**Supplemental Table 3:** Average annual changes in acute coronary syndrome non-fatal incident incidence from 1996 to 2021. Incidence rate trends were analyzed using a Poisson or negative binomial regression model. The rate changes are presented as average annual change percentages during the study period. P-values represent the significance of the trend by study year.

| Incidence |  |  |  |
| --- | --- | --- | --- |
| Age group | Average annual change (%) | 95% CI | P-value |
| **Men** |  |  |  |
| All (35-74)* | -1.16 | -1.37 to -0.95 | 5.70e-27 |
| 65-74* | -2.35 | -2.55 to -2.16 | 3.56e-123 |
| 55-64* | -1.25 | -1.44 to -1.06 | 1.09e-35 |
| 45-54* | -0.47 | -0.67 to -0.26 | 6.26e-6 |
| 35-44 | -0.44 | -0.77 to -0.12 | 0.008 |
| **Women** |  |  |  |
| All (35-74)* | -0.92 | -1.11 to -0.71 | 1.16e-18 |
| 65-74* | -2.48 | -2.69 to -2.27 | 6.27e-117 |
| 55-64* | -0.97 | -1.24 to -0.71 | 1.36e-12 |
| 45-54 | 0.96 | 0.64 to 1.29 | 5.65e-9 |
| 35-44 | 1.57 | 0.86 to 2.29 | 1.35e-5 |

\*Negative binomial model was used instead of Poisson model.

**Supplemental Figure 1:**
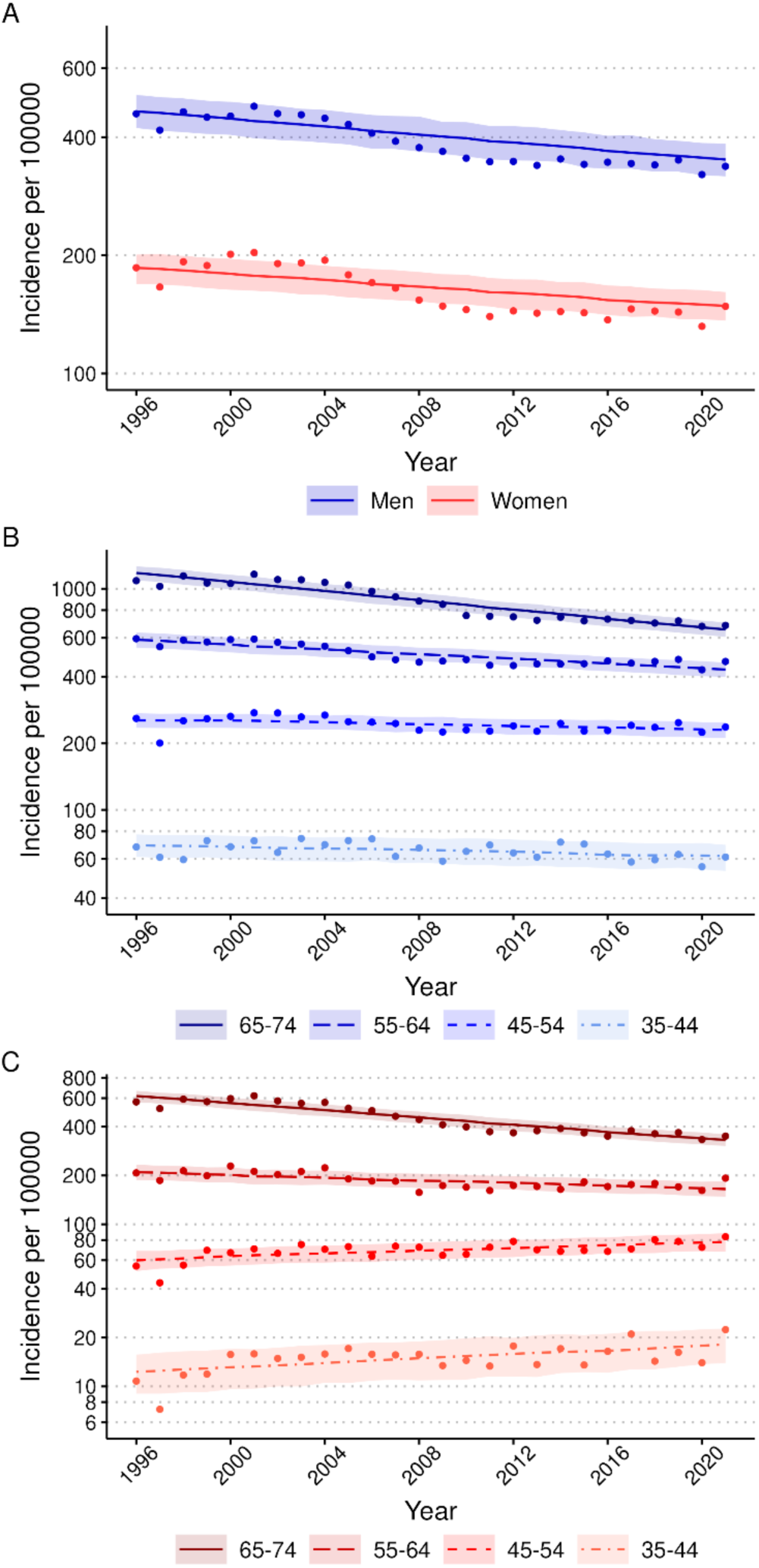
Trends in incidence of first non-fatal ACS incidents in men and women by age group, 1996-2021. A. Incidence rates of men and women aged 35-74. B. Incidence rates of men in 10-year age groups. C. Incidence rates of women in 10-year age groups. Incidence rates include first non-fatal I20.0, I21, and I22 cases. Age-standardized incidence rates per 100 000 inhabitants were calculated with the 2011-2030 European standard population as the reference. Observed incidences are presented as dots, the segmented (Poisson or negative binomial) regression model predicted values as a line and the segmented regression model 95%-confidence intervals for predictions as a ribbon. The rates are presented on a logarithmic scale.

**Supplemental Figure 2:**
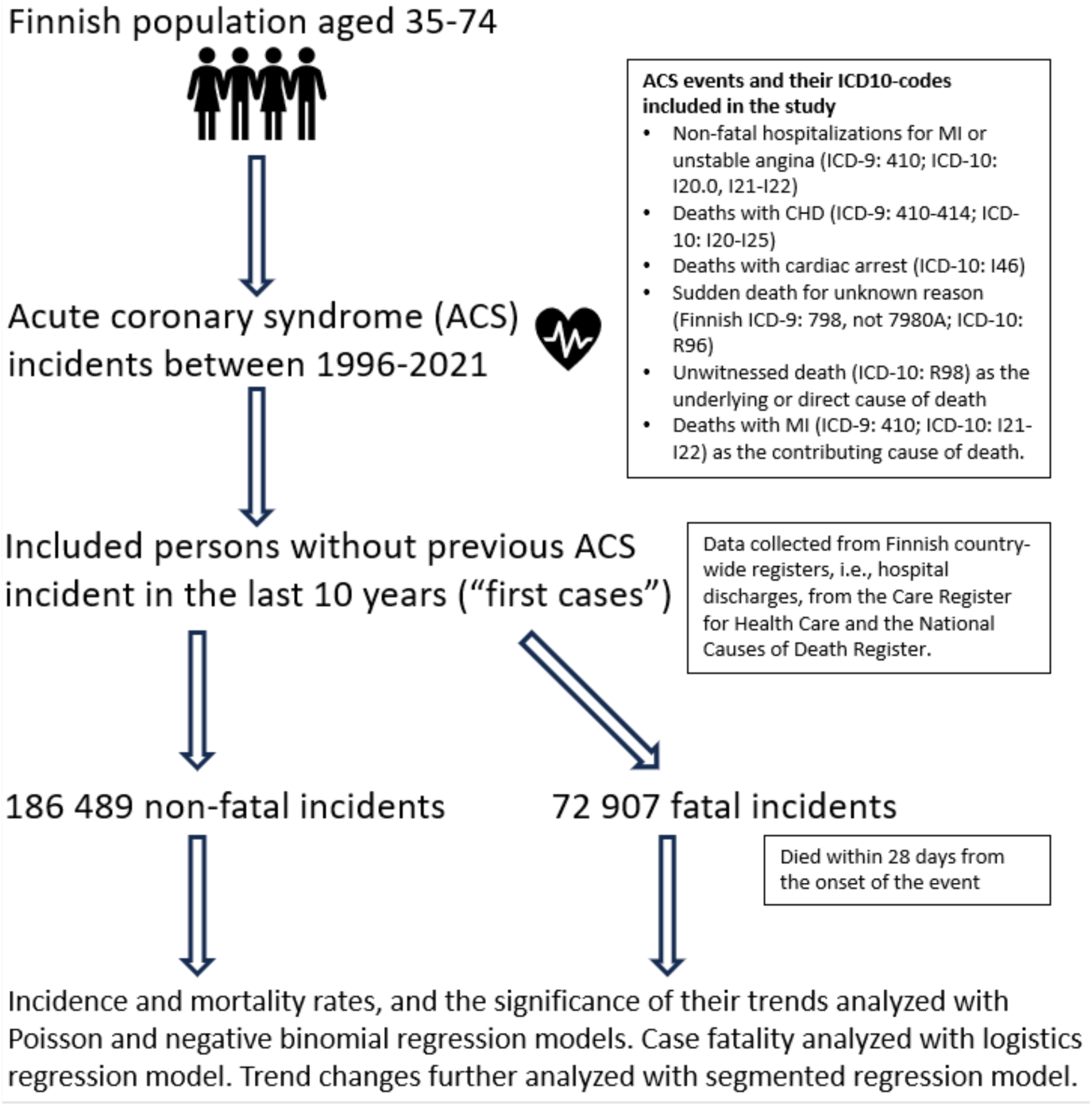
Flowchart of the study population and methods.

**Supplemental Figure 3:**
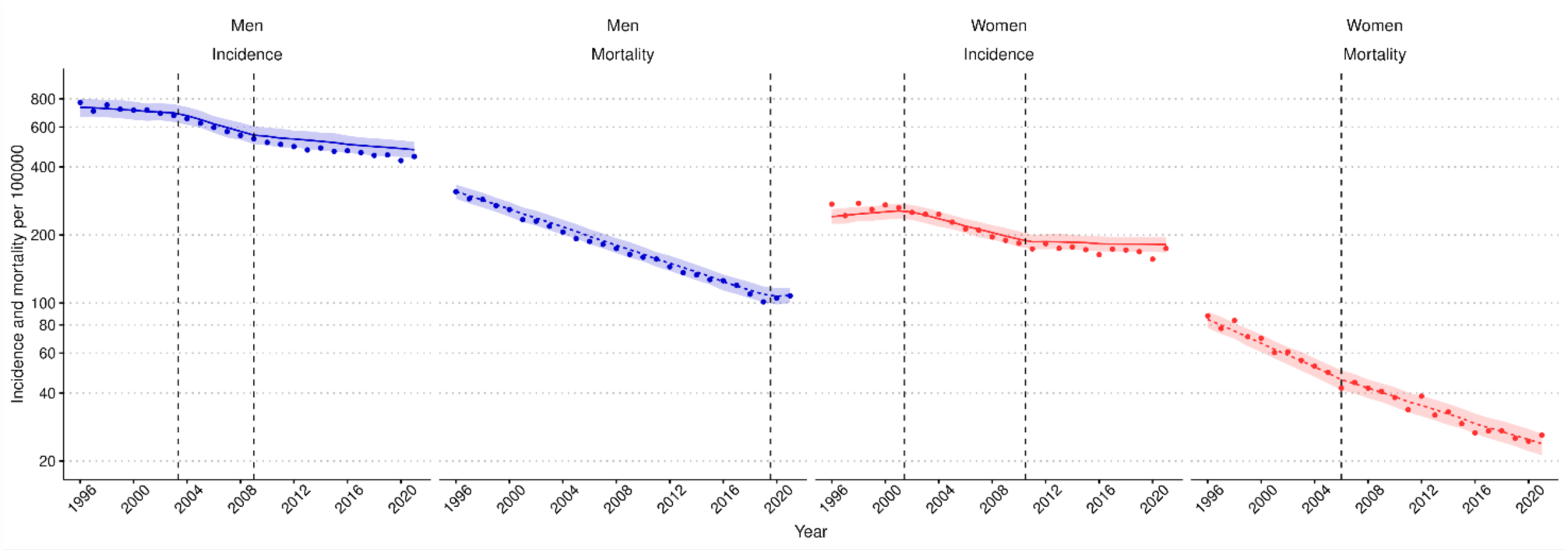
Trends in incidence and mortality rates of acute coronary syndrome in men and women aged 35-75 years, 1996-2021. Rates include first non-fatal I20.0, I21, and I22 cases and fatal I20-25, I46, R96 and, R98 cases. Age-standardized rates per 100 000 inhabitants were calculated with the 2013 European standard population as the reference. Observed incidence and mortality rates are presented as dots, the segmented (negative binomial) regression model’s predicted values as a line, and the regression model’s 95%-confidence intervals for predictions as a ribbon. Breakpoints where there is significant trend change are marked as vertical dashed lines. The rates are presented on a logarithmic scale.

**Supplemental Figure 4:**
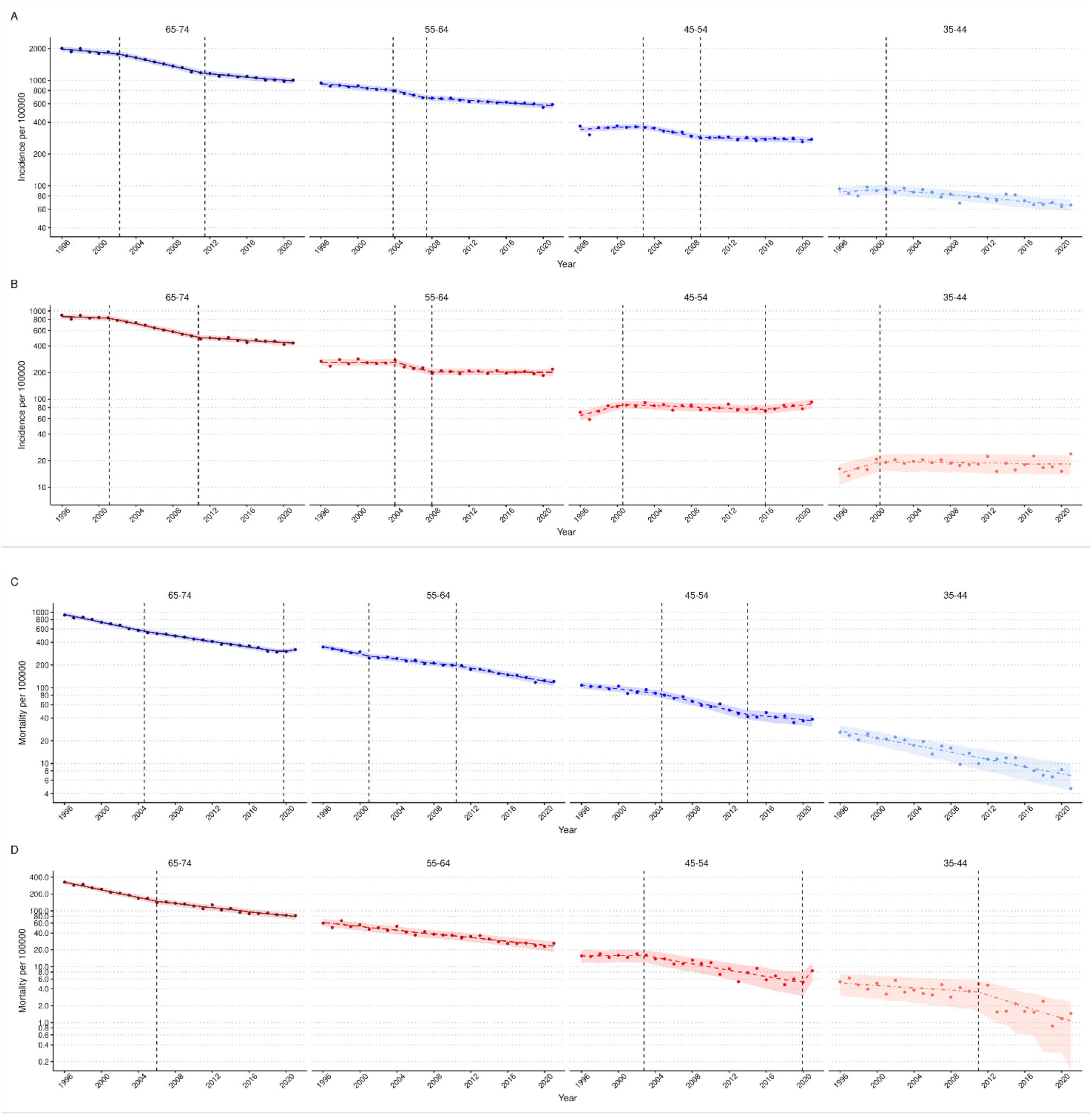
Trends in incidence and mortality rates of acute coronary syndrome in men and women by age group, 1996-2021. A) Incidence rates of men, and B) women. C) Mortality rates of men, and D) women. Incidence rates include first non-fatal I20.0, I21, and I22 and fatal I20-25, I46, R96, and R98 cases. Mortality rates include the fatal cases as mentioned above. Age-standardized rates per 100 000 inhabitants were calculated with the 2011-2030 European standard population as the reference. Observed incidence and mortality rates are presented as dots, the segmented, Poisson or negative regression model’s predicted values as a line, and the regression model’s 95%-confidence intervals for predictions as a ribbon. Breakpoints where there is significant trend change are marked as vertical dashed lines. The rates are presented on a logarithmic scale.

**Supplemental Figure 5:**
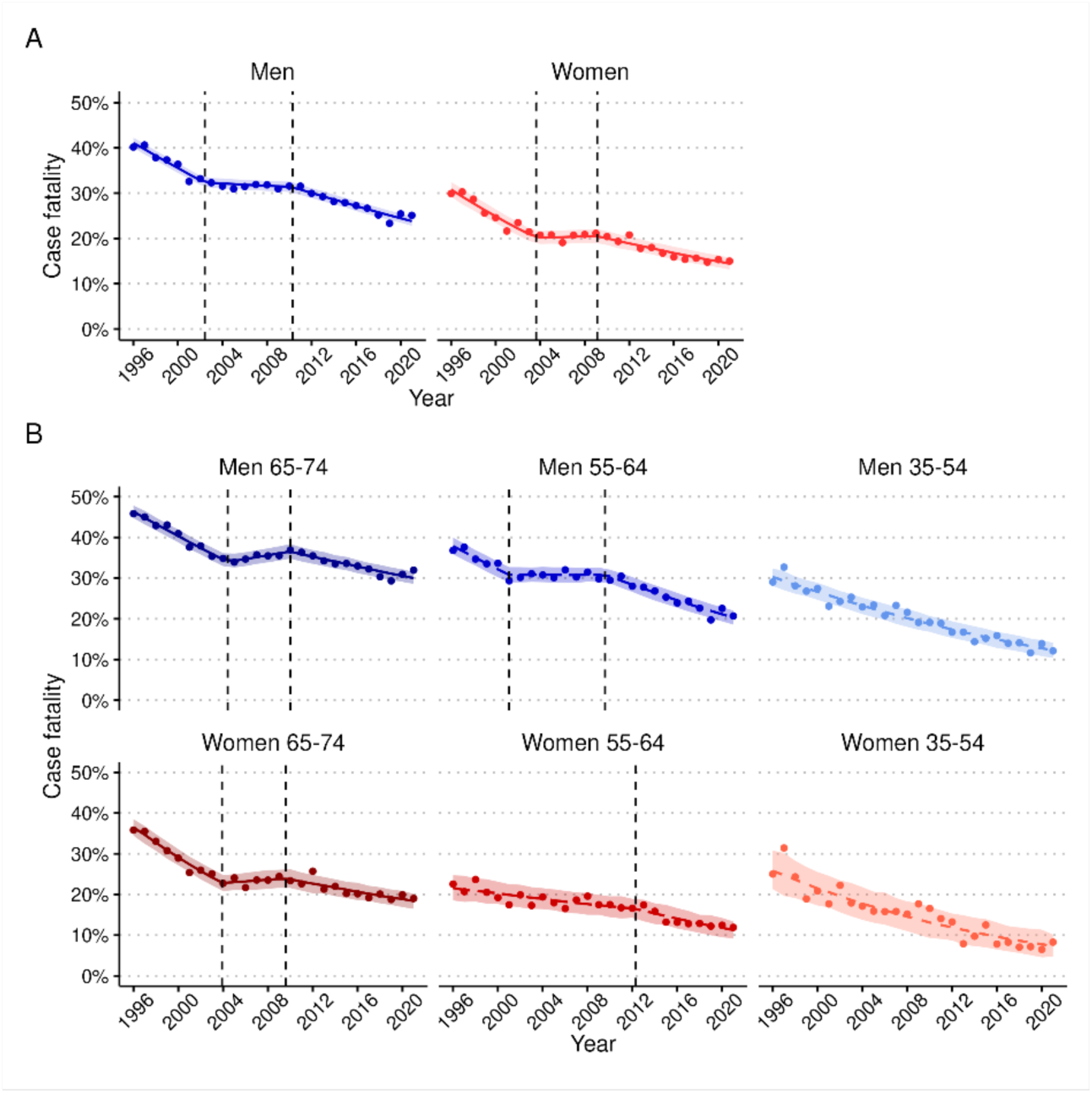
Trends in case fatality (CF) of acute coronary syndrome A) among men and women aged 35-74, and B) by 10-year age groups, 1996-2021. CF includes the first non-fatal I20.0, I21, and I22 and fatal I20-25, I46, R96, and R98 cases. CF was age-standardized using weights based on the age distribution of observed coronary events in populations participating in the WHO-MONICA project. Observed case-fatality is presented as dots, the segmented or logistic regression model’s predicted values as a line, and the regression model’s 95%-confidence intervals for predictions as a ribbon. Breakpoints where there is significant trend change are marked as vertical dashed lines.

